# Imprints of chronic pain trajectories on brain volume and tissue properties

**DOI:** 10.64898/2026.09.18.26362745

**Authors:** Giada Dirupo, Ferath Kherif, Antoine Lutti, Peter Vollenweider, Martin Preisig, Julien Vaucher, Pedro Marques-Vidal, Marc Suter, Chantal Berna, Bogdan Draganski

## Abstract

Despite advances in understanding the mechanistic underpinnings of chronic pain (CP) conditions, knowledge on the brain anatomy microstructure associated to CP trajectories is still scarce. We sought to fill this knowledge gap by combining longitudinal self-reported CP assessments with brain imaging in a community dwelling adult population (n = 976). Aiming to provide anatomical insights beyond morphometry (based on T1 brain images), we analyzed the derived relaxometry measures MTsat, R1 and R2* and we could investigate myelin and iron content. Participants were divided in three groups according to persistent, resolved or new CP and were compared to those without CP at timepoint. Participants with persistent CP exhibited lower grey matter (GM) in fronto-temporal cortical regions and reduced myelin-related signal in subcortical and thalamic areas, alongside higher iron-sensitive metrics in the entorhinal cortex. Those who recovered from CP showed reduced cortical GM and myelin encompassing subcortical nuclei, thalamus, frontal, parietal, and temporal regions. New-onset CP participants displayed reduced GM in cortical areas. Overall, we observed reduced GM volume in a widespread network of fronto-temporal cortical areas in participants with current, resolved and new-onset CP. Myelin-related metrics were lower in the subcortical nuclei and thalamus in participants with persistent and -to a greater extent-with resolved CP, whilst persistent CP was associated with higher iron-sensitive metrics in the entorhinal cortex. Hence, CP trajectories appear to leave differential imprints on brain morphometry and tissue microstructure, with each trajectory associated with distinct non-overlapping spatial and temporal patterns in GM, myelin, and iron content.

## Introduction

Nearly one fifth of the world population suffers from chronic pain (CP) conditions that have significant impact on the individuals’ wellbeing and quality of life[21]. Results from large-scale epidemiological studies brought empirical evidence about the genetic, ethnic and socio-demographic aspects of CP and showed their strong link with morbidity and mortality[22,26,38,53]. Despite major progress in understanding the central nervous system processes related to the persistence of pain, we still have limited knowledge about the brain anatomical correlates of pain trajectories over time.

Computational anatomy using automated feature extraction and statistical analysis of magnetic resonance imaging (MRI) data provides a set of tools for studying the brain anatomy correlates of many conditions including chronic pain. Accumulating evidence points at structural alterations within brain networks implicated in pain anticipation and descending modulation contributing to CP development and persistence[39,41,61,64]. However, findings vary in terms of the specific regions affected and their consistency across chronic pain conditions. Some studies report grey matter (GM) volume reductions in the anterior cingulate cortex (ACC) and anterior insula (AI), see[61] for a meta-analysis. Other studies demonstrate the key role of the dorsolateral prefrontal cortex (DLPFC), with results from a narrative review highlighting reduced GM volume in participants with CP[41]. Longitudinal studies went further to demonstrate the association between brain structural changes and CP persistence, with GM volume reduction in cortical regions both in recent and persistent CP, but only persistent CP showing reduction in subcortical structures[66]. Furthermore, resolved CP has a transitory character evidenced by a reversal of the relative volume reduction once pain is resolved[11,19].

There is also growing evidence that complex interactions among sensory and memory consolidation mechanisms contribute to the development and persistence of CP, with theoretical and empirical work putting forward the idea of CP arising from dysfunctional cortico-hippocampal circuits related memory consolidation[40,43,44]. This notion is further supported by evidence of entorhinal involvement both in pain anticipation processes in healthy controls[17,52] and a reduced GM volume in CP patients[43]. The neuroanatomical basis for CP onset and persistence and its interactions with sensory memory systems in complex, time-dependent ways-remains without conclusive evidence.

While these patterns suggest some shared neurobiological features between chronic pain conditions, studies diverge on regional specificity, temporal dynamics, and brain microstructural characteristics beyond gray matter volume. Besides clinical, methodological, and analytical differences, conflicting results may reflect the field’s reliance on T1-weighted imaging, which risks spurious morphometric findings[33] and might reflect processes other than grey matter volume thickness, such as GABA_A_ receptor density[50]. Relaxometry-based quantitative MRI (qMRI) holds the promise to minimize this risk by providing higher sensitivity to the unique contribution of brain’s microstructural components and allowing for an attempt at a neurobiological interpretation of the obtained results. Despite its growing application across clinical conditions and population-based studies[5,34,57], to date neural qMRI in CP has only been employed in one study, which found differences in iron levels in subregions of the cerebellar, insular and frontal lobe in patients with headaches related CP[8].

Chronic pain involves dynamic histological remodeling across nociceptive circuits, including structural changes in peripheral sensory neurons, synaptic reorganization within the spinal dorsal horn, and alterations of cortical sensory maps and brain connectivity[4,28]. These processes are increasingly linked to neuron–glia–immune interactions and neuroinflammatory mechanisms, which appear to contribute to the maintenance of persistent pain and show regionally specific associations with pain characteristics[12,32].

Supporting the notion of a wide variation in CP trajectories, a recent study confirmed the importance of socio-demographic and general health related factors as predictors of pain persistence and recovery[14]. Here, we sought to use a linked MRI database to provide empirical evidence for the association between individuals’ CP trajectories and brain anatomy focusing on the detailed investigation of tissue microstructure (markers of myelin and iron contents) alongside established morphometry. We leveraged the statistical and interpretational power of longitudinal pain assessments and cross-sectional qMRI data (n>800) in a cohort of community-dwelling individuals. Our hypothesis was that individual CP trajectories (persistent, resolved and developed) would show an association with brain circuits involved in pain downregulation/anticipation and memory consolidation.

## Methods

### Study participants

The data stems from the BrainLaus (https://www.colaus-psycolaus.ch/ professionals/brainlaus/), part of the CoLaus|PsyCoLaus longitudinal cohort including at baseline 6’734 subjects aged from 35 to 75 years from the community-dwelling population living in the city of Lausanne, Switzerland. Participants were recruited between 2003 and 2006 and had physical and psychiatric baseline evaluations between 2003 and 2009 [18,51]. A physical and psychiatric follow-up (FU1) took place between 2010 and 2014 and a second (FU2) between 2014 and 2018. Subjective pain reports using structured questionnaires were introduced at the FU1 and MRI for the BrainLaus study at the FU2. For the present paper we used the subjective pain reports of FU1 and FU2 (see [14]) together with the MRI data acquired at FU2 (n = 976; 512 women, age = 59.94 ± 9.11 years old), see below and [34] for details on data acquisition and preprocessing. All participants with available data at both FUs for self-reported pain and at FU2 for brain structural data were included in the study, see Results section for details.

The institutional Ethics Committee approved the CoLaus|PsyColaus baseline investigation (reference 16/03; 187-03, 134-05bis, 134-05-2 to 5 addenda 1 to 3), the first (reference 33/09; 239/09), and the second (reference 26/14; 239/09 addendum 1 and 2) follow-up. All participants signed a written informed consent prior to their participation. The study was performed in agreement with the Helsinki declaration, and in accordance with the applicable Swiss legislation.

### Pain assessment

We defined chronic pain as daily pain lasting at least 3 months, a criterion based on the ICD-11 definition [63], and operationalized by as a positive answer to both screening questions: ‘Do you currently suffer from pain on a daily basis in one or multiple locations?’ (Y/N) and ‘Since when you suffer from daily pain?’ (>3 months Y/N) [3,14]. This combination of factors led to a variable of “chronic pain” (present/absent; CP +/-) at each follow up.

Aiming to differentiate the short and long-term effects of CP on the brain, we stratified study participants according to a cross-sectional and trajectory-based definition [14]. We first determined the presence (CP+) or absence (CP-) of CP at FU2 as criterion for studying the brain-CP associations at that time point. We then divided the sample into four subgroups depending on the presence or absence of CP at FU1 and FU2 allowing us to investigate the persistence or improvement of CP between the two timepoints. The four possible combinations were represented respectively by: a group of participants that did not report CP in either of the FUs: the “never CP” group. A second group, “persistent CP” which included those who reported pain at both follow-ups. Those who had CP only at FU2 (“developed CP”) and finally the “recovered CP” group consisting of all participants who reported CP at FU1 but not at FU2 [14].

### MRI data acquisition and pre-processing

For our study we analysed the qMRI data acquired for the BrainLaus nested study on a 3T whole-body machine (Magnetom Prisma, Siemens Medical Systems, Germany), using a 64-channel radio-frequency (RF) head coil and body coil for transmission. The qMRI protocol consisted of three multi-echo 3D fast low angle shot acquisitions with magnetization transfer-weighted (MTw: TR = 24.5 ms, α = 6°), proton density-weighted (PDw: TR = 24.5 ms, α = 6°), and T1-weighted (TR = 24.5 ms, α = 21°) contrasts with 1mm3 isotropic resolution [23–25]. To correct for the effects of RF transmit field inhomogeneities, we acquired B0 (2D double-echo FLASH sequence with slice thickness = 2 mm, TR = 1020 ms, TE1/TE2 = 10/12.46 ms, α = 90°, BW = 260 Hz/pixel) and B1 maps (3D echo-planar spin-echo and stimulated echo images with 4 mm3 resolution, TE = 39.06 ms, TR = 500 ms) [36,37]. The total acquisition time was 27 min.

For calculation of the magnetization transfer saturation (MTsat), transverse relaxation rate (R2* = 1/T2*), effective longitudinal relaxation rate (R1 = 1/T1) and effective proton density (PD*) maps we used the voxel-based quantification (VBQ) hMRI toolbox implementation [16,55] within the SPM12 framework (Wellcome Centre for Human Neuroimaging, London, UK). First, we calculated probabilistic maps of grey matter (GM), white matter (WM) and cerebro-spinal fluid (CSF) from MTsat and PD* images using the multi-channel “unified segmentation” and enhanced tissue priors [33]. This was followed by an atlas-based parcellation of GM structures using factorisation-based labelling [2] and calculation of regional averages of tissue volume, MTsat, R2* and R1. Aiming at higher granularity of thalamic, hippocampus and periaqueductal grey substructures, we used additional labels according to previously published definitions [27,30,60]. Finally, we estimated a proxy for individuals’ head size - the total intracranial volume (TIV) by summing the GM, WM and CSF volumes. Similarly, across parameter maps, we add all GM regional MT, R1, R2* and PD* values to provide corresponding global whole-brain estimates.

### Statistical Analyses

For statistical comparison of the CP-stratified groups we used chi-squared and t-tests. The analysis of the brain imaging data was performed using linear multiple regressions where single ROI-based estimate was regressed independently against CP (presence/absence at FU2) including age, gender and corresponding global metric (TIV or MTsat, R1 or R2*) as a priori selected covariates of no-interest, following well-established modeling approaches previously applied to this dataset. [20,57]. We repeated this analysis for each parameter map to test for group differences across ROIs and used Bonferroni corrections for multiple comparisons. To analyse brain tissue properties of CP over time, we used an analogous model but regressed each ROI against the CP trajectories variable. When a significant effect was detected, post hoc pairwise comparisons between trajectory groups were conducted using Tukey Honest Significant Difference test for each pair of groups, which controls the family-wise error rate within the set of pairwise contrasts. We report results at significance level p<.05, after family-wise error (FWE) correction for multiple comparisons. Trends are reported at p<.001, uncorrected for multiple comparisons.

### Quality assessment

The degradation of the MRI data due to participants’ head motion was assessed using the validated motion degradation index (MDI) described in [6,35]. Separate MDI values were computed for the MT, T1 and PD-weighted images, used for the computation of the relaxometry maps [10,35]. MDI values from the MT-weighted images are higher than those from the PD- and T1-weighted images by ∼1s-1 due to the lower number of echo images and lower signal-to-noise of the raw data (Lutti et al., 2022). Although datasets with MDI values below 6s-1 for the PD- and T1-weighted images, and 7s-1 for the MT-weighted images are empirically considered of sufficient quality [6,35], for the present study, we set a conservative threshold of 5s-1. We chose to apply this single, common threshold across all three maps in order to minimize data loss while keeping the level of accepted motion degradation comparable to that of prior studies [35]. We thus excluded from the analyses all subjects with scans above this cutoff, resulting in a final sample of 837 subjects.

## Results

### Participants’ stratification

The stratification according to CP at FU2 resulted in two groups with 334 CP+ individuals (204 females, mean age 58.6 ± 8.4 years old), and 503 CP- (248 females, mean age 58.6 ± 9.0 years old). The two groups did not show significant differences in age (t_(773.15)_= -0.02, *p*= 0.981), while more participants in the CP group were females (χ^2^= 8.49; *p*= .004), (See Figure 1).

**Figure 1:**
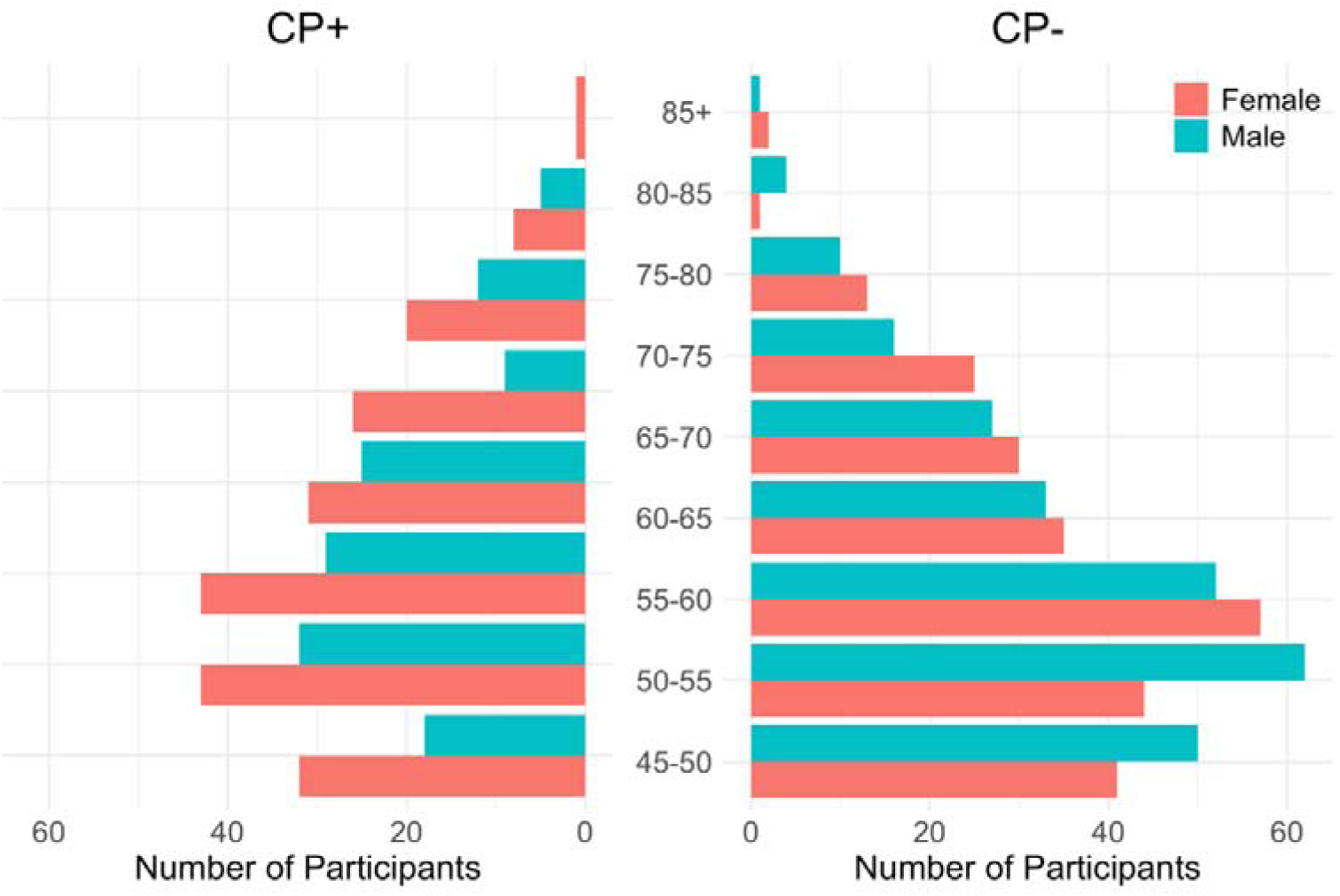
Gender and age distribution of the sample. Pyramid graph showing the distribution of participants in the CP+ (left) and CP- (right) sub-samples at FU2. All participants included in the final analyses are included (i.e. those who both responded on the CP self-reported questionnaire and underwent MRI scan which passed the quality assessment). On the x-axis the number of participants per gender (women=red, men=blue) and age range. On the y-axis the age, ranked in categories of 5 years from 45 years.

The stratification according to CP trajectories generated the four subgroups: i. “never CP” (n=382, 190 females, aged 58.34y ± 8.96); ii. “persistent CP” (n= 196, 127 females, aged 58.72y ± 8.37); iii. “developed CP” (n=117, 56 females, aged 59.67y ± 9.15); iv. “recovered CP” (n=131, 74 females, aged 58.24y ± 8.55). These groups did not differ in age (ts< -1.39; *p*s> 0.165). Chi-squared tests on each pair of groups showed an imbalance in the subjects’ gender proportion with the “persistent CP” group counting more female participants as compared to the “never CP” (χ^2^= 11.26; *p*<.001) and the “developed CP” (χ^2^=7.97; *p*= 0.004) subsamples.

### Cross-sectional analysis

The comparison between CP+ and CP- individuals at FU2 did not show any significant results for GM volume, MT, R1 or R2*. We denote the trends for lower GM volume in CP+ patients in the nucleus accumbens (NAcc), thalamus, frontal and temporal lobes, lower MTsat in the thalamus, higher R2* values in hypothalamus, entorhinal cortex and lower R2* values in the PAG (see Table 1S Suppl. material).

### Longitudinal analysis

When comparing the four subgroups defined by the CP trajectories, we observed a widespread pattern of MT, R1 and R2* differences in subcortical areas, whilst the cortical areas - e.g. temporal cortex, showed predominantly volume and MTsat differences (See figure 2).

**Figure 2.**
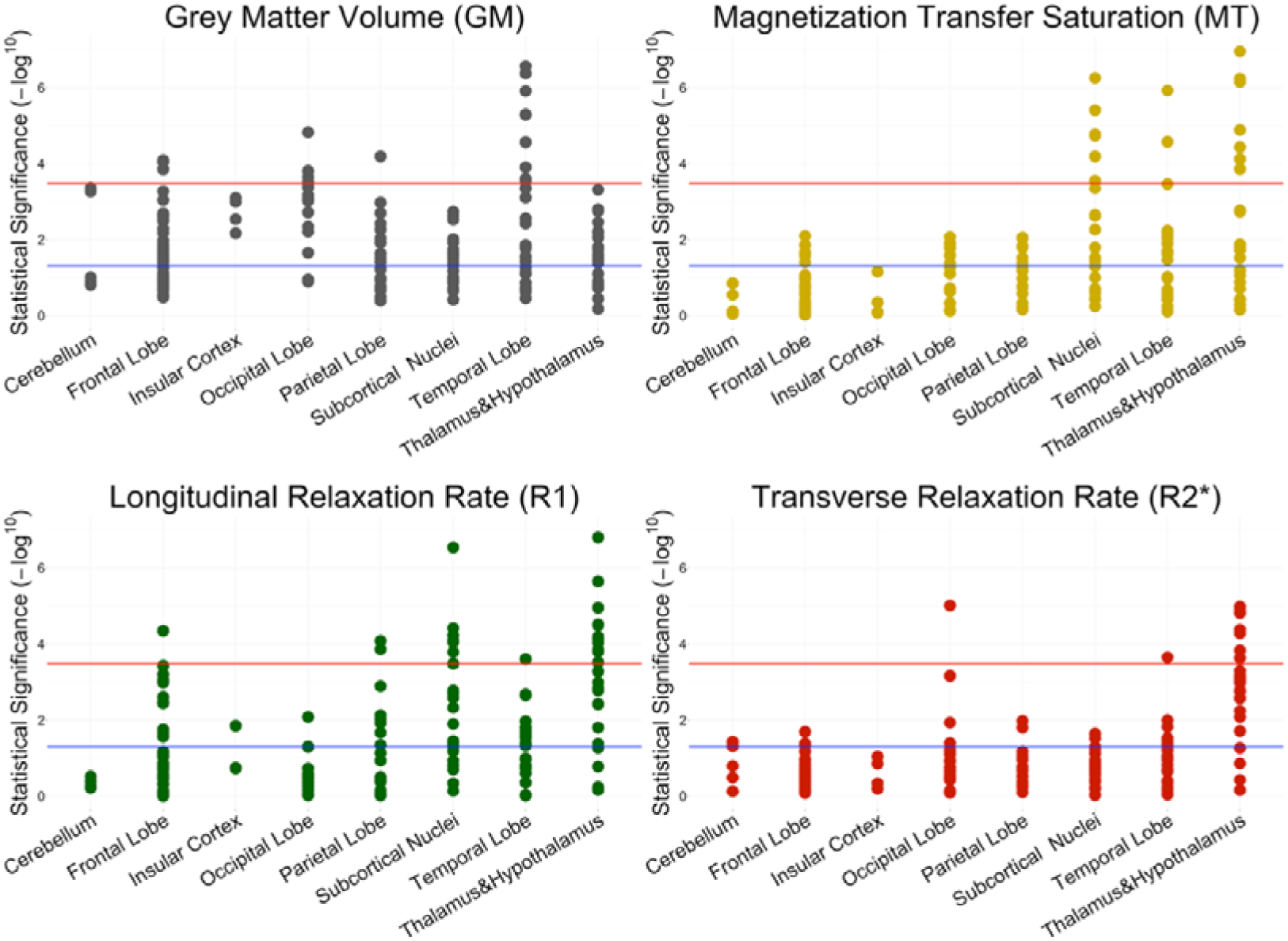
Manhattan plot of brain anatomy differences according to CP Trajectories. Each dot represents a model where an ROI was regressed against CP trajectory. On the y-axes, is the –log10 of the p-value of that model, where a higher value is associated to lower p-value (statistical significance level). The blue lines represent the .05 alpha level and red lines represent the Bonferroni correction cut off.

For all between-group comparisons, we considered the “never CP” group as the reference (see Figures 3 and Table S2 and S3). The post hoc analyses showed that the “developed CP” group differed the least from the other groups in terms of microstructural changes, indeed we found higher R1 values in the putamen bilaterally and lower R2* in the fusiform gyrus bilaterally (see Figure 3, DCP bars across all panels). The GM volume was smaller in the left frontal operculum and posterior orbital gyrus and right medial orbital gyrus, as well as in the right inferior occipital gyrus, occipital pole and superior occipital gyrus. Finally, in the temporal lobe many ROIs showed reduced volume in comparison to the baseline, namely in the entorhinal cortex, inferior temporal gyrus, temporal pole bilaterally and the right hippocampus (see tableS2).

**Figure 3:**
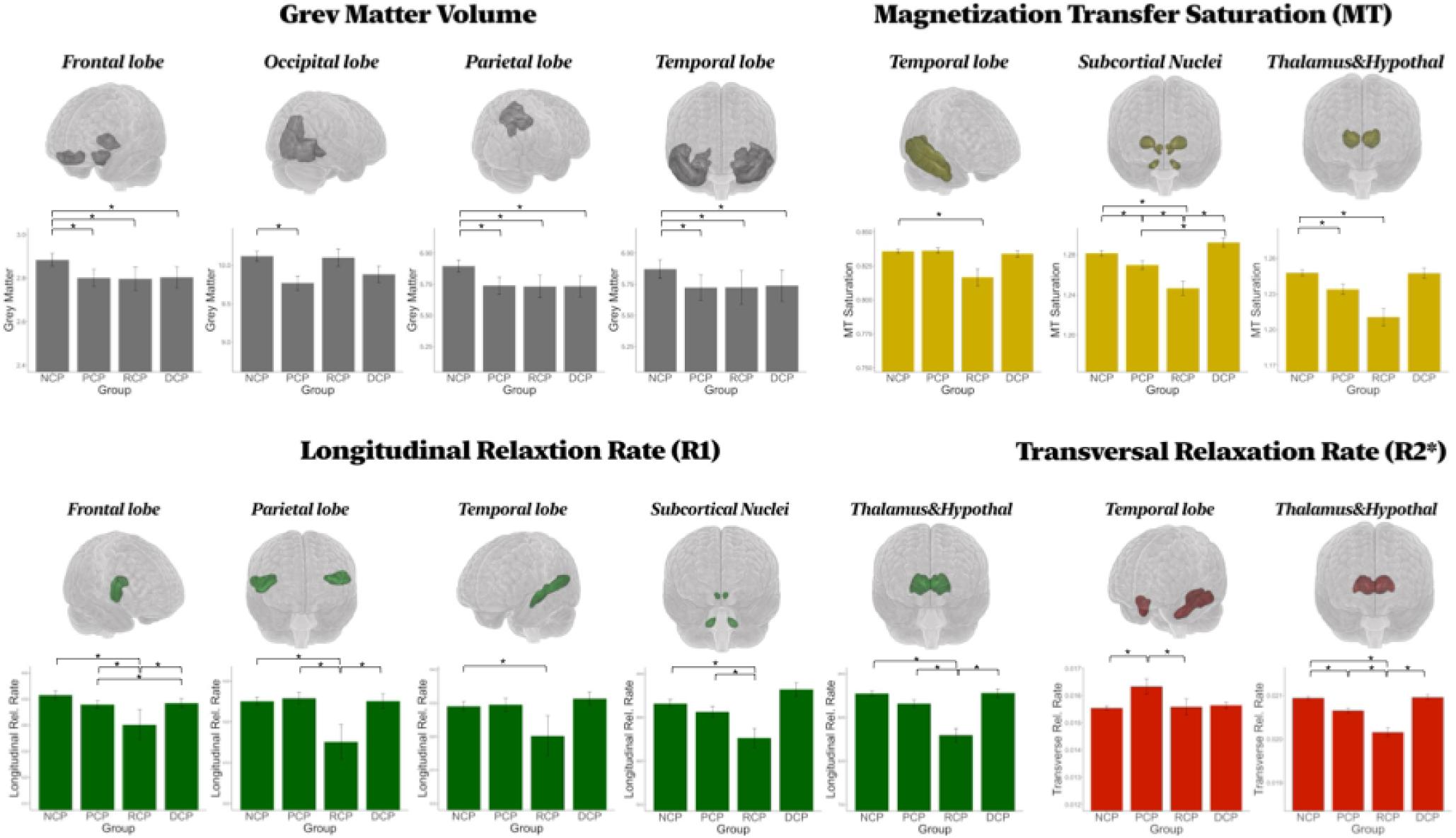
Brain Anatomy metrics associated to the CP Trajectories. Each brain map and related bar plot under it refer to a lobe of the brain (grouped as in Figure 2) and show an ROI -or a group of ROIs-with significant differences across CP Trajectories. The four histograms refer to the four CP trajectories (NCP= never CP, PCP= persistent CP, RCP= recovered CP, DCP= developed CP) and highlight the direction of the effect. Significant post-hoc results are highlighted by a *.

The group of participants who reported persistent CP (see Figure 3, PCP bars across all panels) showed lower MTsat in the medial dorsal thalamus bilaterally and right deep cerebellar nuclei. We also observed lower R2* in the anterior thalamus bilaterally contrasted with higher R2* in the entorhinal area bilaterally. The left frontal operculum, posterior orbital gyrus and medial orbital cortex, in the right inferior and superior occipital gyrus, lingual gyrus and occipital pole and in the superior parietal lobule showed GM volume decreases. Similar GM reduction was observed in the parahippocampal gyrus, the entorhinal cortex, inferior temporal gyrus, temporal pole bilaterally, the right hippocampus (see table S2).

The “recovered CP” group (see Figure 3, RCP bars across all panels) showed the most widespread pattern of MTsat and R1 differences confined to the right deep cerebellar nuclei, bilateral red nuclei, bilateral pallidum and left nucleus tuberomammillary of the hypothalamus, in the bilateral thalamus (medial dorsal, VL. VP) and the superior temporal gyrus. There was an MTsat decrease in the right substantia nigra, the subthalamic nucleus and the right temporal gyrus (middle portions). We report decreased R1 in the right IFG, bilateral putamen, portions of the thalamus (right intralaminar, bilateral pulvinar nucleus, bilateral VA and PA) and finally in the bilateral central operculum and right operculum of the parietal lobe. We observed lower R2* in the right intralaminar, bilateral VA, A and mediodorsal thalamus and the fusiform gyrus bilaterally. Similarly, there was GM volume reduction in the right medial and left posterior orbital gyrus, the right inferior and superior gyrus and occipital pole occipital lobe, the parahippocampal gyrus, entorhinal area, inferior temporal gyrus, temporal pole bilaterally and the right hippocampus.

## Discussion

In this study we demonstrated patterns of brain morphometry and tissue property differences associated to CP trajectories spanning over 10 years of observation. The reported neuroanatomical findings related to dynamic changes in pain status were opposed to a lack of significant differences in the cross-sectional analysis. The CP microstructural characteristics confined to the thalamus, basal ganglia, frontal, temporal and parietal lobe structures were predominantly symmetrical and characterized by differences in indices sensitive to myelin and iron content. We interpret these results, though obtained from cross-sectional imaging data, as suggestive for dynamic processes that begin with the occurrence of CP and evolve over time depending on the trajectory in and out of pain or to its persistency.

When comparing the four CP trajectories, we observed a distinctive pattern of changes over time in both volume and brain tissue microstructure, with marginal spatial overlap between them except for the entorhinal cortex. This suggests that transitions into and out of CP are associated with brain anatomy characteristics that are regionally specific, with each trajectory associating to volume, myelin, or iron alterations in distinct neural structures. Supporting this interpretation, we found lower GM volume in the cortex across trajectories, while specific group differences in microstructural characteristics were evident in subcortical areas. As R1 values are used as metrics of both myelin an iron content, MTsat of myelin [55], and there was remarkable similarity between MTsat and R1 patterns, we take this as supportive evidence for an interpretation directed towards myelin content.

Individuals with newly developed CP resembled healthy controls the most, particularly in their myelin and iron-related microstructural brain characteristics, suggesting brain microstructure changes slowly after CP onset. Smaller GM volume was detected across frontal, temporal, and parietal regions, consistent with prior research on CP [41,61,66], suggesting this process begins in early chronic pain phases. R1, MT, and R2* maps showed no baseline differences, indicating myelin and iron changes occur later or during recovery. We found that persistent CP was related to a reduction of cortical volume in a widespread network of the frontal, temporal, occipital and parietal areas. Importantly, some of those areas, notably the frontal ones, have been consistently associated to pain modulation (see [41] for an overview), suggesting a possible altered functioning in the case of persistent CP. While previous studies highlighted a smaller volume of the thalamus and subcortical nuclei in CP+ patients, across different CP conditions [29,47,48], we observed changes in the values indicative of myelin and iron content, but not GM. This divergence could be due to prior work misinterpretation of the group difference in thalamus values as a change in volume, while it was myelin and iron content the main contributors of this result [33]. This misinterpretation is in line with findings of disrupted connectivity in tracts surrounding and connecting the thalamus [42] and structural sequences combination (rT1/T2) signal in thalamus and subcortical nuclei in CP+ patients [59]. Finally, the entorhinal cortex showed an increased level of R2* values compared to participants not reporting CP at either FU, suggesting that persistent CP is associated to higher iron content in this area. The entorhinal cortex plays a crucial role in temporal association memory [17,52]. Indeed its activity is associated to tonic (as opposed to phasic) pain in animal models [65] and anticipation -including subsequent modulation of-pain [43]. While iron accumulation in the brain is generally associated to neuroinflammation [54,58,62], it also plays a crucial role in dysfunctional neuronal energy metabolism [49]. However, additional inflammatory markers were not available for analysis in this project, precluding a more direct assessment of whether the observed iron-related brain microstructural changes are associated with inflammatory processes. Inflammation is therefore discussed as a plausible mechanistic interpretation supported by prior literature rather than as a tested covariate in our analyses. Our results highlighted an abnormality in the entorhinal cortex region in individuals with long-term CP, potentially reflecting sustained hyperactivation, neuroinflammatory processes, or altered iron homeostasis [49,54,58,62]. We do not interpret those effects as due to aging or associated to cognitive and memory impairment since our cohort, despite including participants of advanced age, generally did not show significant cognitive limitations. This is confirmed by their MMSE scores (above 29) published recently in another article (see table in [45]. Indeed, this is often the case in epidemiological cohorts where the ageing population is biased by the absence of cognitively impaired individuals, who tend to dropout.

In subjects who recovered from CP, as compared to participants not reporting CP at either FU, we found smaller GM volume in frontal parietal and temporal brain areas, suggesting that GM does not restore within 5 years after recovery. We found no increase of GM, contrary to previous studies that show restored GM after recovery in the frontal cortex (prefrontal, supplementary motor area), and subcortical region (hippocampus, insula, amygdala and thalamus) [11,19], suggesting a possible misattribution of higher t1-weighted signal identified in XYZ area in priori studies [33]. Areas of the cortex (temporal, frontal and parietal), the thalamus and subcortical nuclei showed altered R1 and MT values. We interpret these findings as reflecting the loss of previously established dysfunctional connectivity patterns associated with CP, which may be diminished as individuals transition again toward a pain-free state. Finally, the thalamus also showed lower R2* values, which may be mechanistically linked to the reduced myelin observed here, given that iron is essential for oligodendrocyte-mediated myelin synthesis [7,46,56].

We did not find differences when comparing CP+ and CP-individuals, which is at odds with previous studies [9,13,15,31,41,61]. Besides potential explanations related to sample size and unaccounted selection bias in small-scale studies, the difference in the imaging protocols between our work and prior studies needs to be underscored. In fact, qMRI holds the promise to minimize spurious morphometric findings [33]. Different anatomical distributions of pain [9,15], socio-demographic factors (e.g. based in different geographic regions [31]) or definitions of chronicity of pain [15] could be additional potential explanations for the lack of agreement with previous studies.

## Conclusion

Overall, our findings sustain that CP affects the brain subcortical microstructure and cortical volume in different ways depending on the trajectories in and out of chronic pain and its persistence in time. We demonstrated a change in microstructural metrics associated to structural connectivity among CP trajectories expressed by two different myelin content concentration parameters. We interpret this result as due to plasticity processes such that the observed decrease in MTsat in persistent CP [1] could represent the loss of previously present neural pathways in the thalamic and subcortical regions. In those recovering from CP, we detected the weakest MTsat, possibly indicating that dysfunctional connectivity pathways are no longer present to the same extent (leading to a reduced myelin content), but other alterations could impair a full/ fast recovery of the healthy pathways. Future studies should investigate the long-term consequences of CP after recovery (at >5 years) in terms of brain volumetrics and microstructures. Smaller volumes in the cortical areas were detected in new and persistent CP, as well as after recovery, suggesting that this process is little reversible. Finally, in the persistent phase, the entorhinal cortex was affected in terms of both its morphology and tissue properties, highlighting the crucial role of this brain structure in CP trajectories over time. Taken together, these results suggest that chronic pain is characterized by dynamic, trajectory-dependent alterations in brain morphology and tissue properties. Rather than reflecting stable group differences, the observed changes point to regionally specific processes associated with pain onset, persistence, and recovery, highlighting the value of quantitative MRI approaches for capturing long-term neurobiological reorganization to chronic pain.

### Limitations

While the longitudinal dataset we used constitutes a strength of this study, the precise time of onset and offset of chronic pain between the cohorts’5 years questionnaires are not known. This, combined with a cross-sectional brain data collection at only the second time point of our study is limiting causal and temporal inference. Further studies from the same cohort, including modeling data from further time-points could provide deeper knowledge on correlations between brain morphometric properties and clinical outcomes. Additionally, the data collected from this general population sample did not allow for stratified analyses by specific pain characteristics, including pain mechanism (e.g., neuropathic versus nociceptive), affected body region, or etiological factors, which may have masked subgroup-specific associations.

## Acknowledgements

B.D. is supported by the Swiss National Science Foundation (project grant no. 213595, 32003B_135679, 32003B_159780, 324730_192755 and CRSK-3_190185), ERA_NET NEURON JTC2020: iSEE and JTC2023-ELSA: BrainTree projects. A.L. is supported by the Swiss National Science Foundation (grant no. 320030_184784, CR00I5-235940). The Laboratory for Research in Neuroimaging (LREN) is very grateful to the Roger De Spoelberch and Partridge Foundations for their generous financial support. F.K is supported by the PHASE IV AI project (agreement no: 101095384) funded by the European Commission through the Horizon Europe program.

The CoLaus study was supported by unrestricted research grants from GlaxoSmithKline, the Faculty of Biology and Medicine of Lausanne, the Swiss National Science Foundation (grants 3200B0– 105993, 3200B0-118308, 33CSCO-122661, 33CS30-139468, 33CS30-148401, 33CS30_177535, 324730_204523, 320030_220190) and the Swiss Personalized Health Network (grant 2018DRI01).

## Data availability statement

The data of CoLaus|PsyCoLaus study used in this article cannot be fully shared as they contain potentially sensitive personal information on participants. According to the Ethics Committee for Research of the Canton of Vaud, sharing these data would be a violation of the Swiss legislation with respect to privacy protection. However, coded individual-level data that do not allow researchers to identify participants are available upon request to researchers who meet the criteria for data sharing of the CoLaus|PsyCoLaus Datacenter (CHUV, Lausanne, Switzerland). Any researcher affiliated to a public or private research institution who complies with the CoLaus|PsyCoLaus standards can submit a research application to. Proposals will be evaluated by the Scientific Committee (SC) of the CoLaus|PsyCoLaus study. Detailed instructions for gaining access to the CoLaus|PsyCoLaus data used in this study are available at www.colaus-psycolaus.ch/professionals/how-to-collaborate/.

## Supplementary Material

**Table S1.** Uncorrected results from the model where the presence of CP and was regressed against each ROI after adjusting for age, sex, and total intracranial volume (or sum of compound of interest). only p-values of the significant results from the uncorrected model are shown for each biophysical model.

| ROI | GM | MT | R1 | R2* | PD |
| --- | --- | --- | --- | --- | --- |
| r/l Accumbens | 0.009 |  |  |  | 0.036/0.045 |
| r Calcarine Cortex | 0.017 |  |  |  |  |
| r/l Entorhinal Area |  |  |  | 0.001/0.043 |  |
|  | 0.026/0.047 |  |  |  |  |
| l Frontal Operculum | 0.016 |  |  | 0.042 |  |
| r/l Medial Orbital Gyrus | 0.018/0.037 |  |  |  |  |
| r/l Occipital Pole | 0.019 |  |  |  | 0.021/0.043 |
| r Posterior Orbital Gyrus | 0.049 |  |  |  |  |
| r Superior Parietal Lobule | 0.001 |  |  |  |  |
| r Thalamus (anterior) | 0.005 |  |  |  |  |
| r Thalamus (intra laminar) | 0.018 | 0.022 |  |  |  |
| l Thalamus (ventro anterior) | 0.032 |  |  |  |  |
| r Anterior Thalamus (posterior) | 0.032 |  |  |  |  |
| r Anterior Insula |  |  | 0.043 |  |  |
| r/l Hypothalamus (nucleus tuberomam) |  |  | 0.006/0.023 | 0.007/0.002 | 0.012/0.022 |
| r Cuneus |  |  |  |  | 0.042 |
| r Planum Polare |  |  |  |  | 0.032 |
| r Planum Temporale |  |  |  |  | 0.015 |
| r Transverse Temporal Gyrus |  |  |  |  | 0.004 |
| r Anterior Hypothalamus |  |  |  | 0.011 |  |
| l Inferior Frontal Gyrus (orbital part) |  |  |  | 0.049 |  |
| r Precentral Gyrus |  |  |  | 0.047 |  |
| PAG |  |  |  | 0.013 |  |

**Table S2 and S3.**
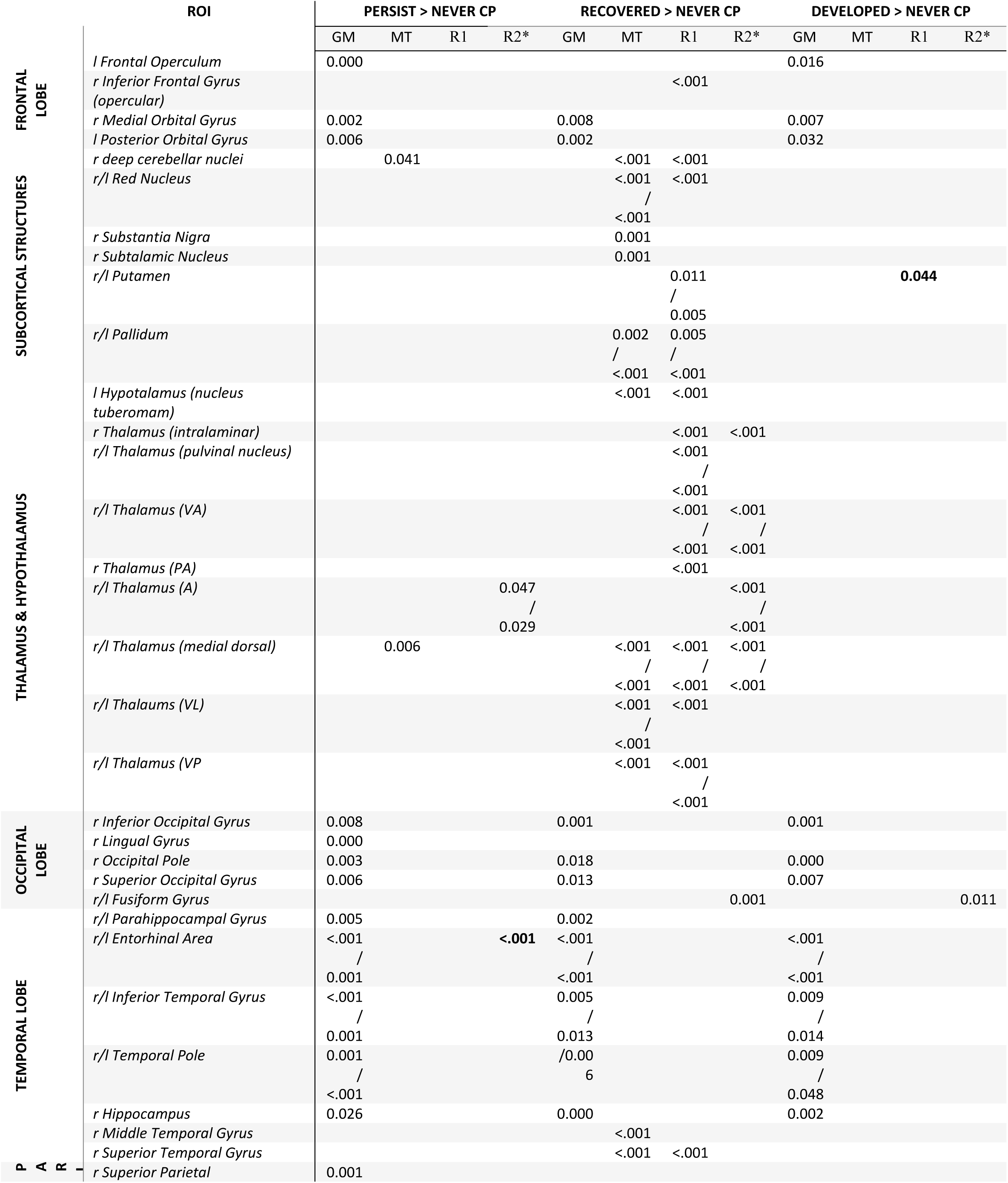
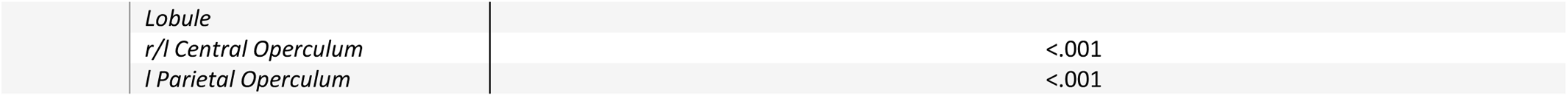
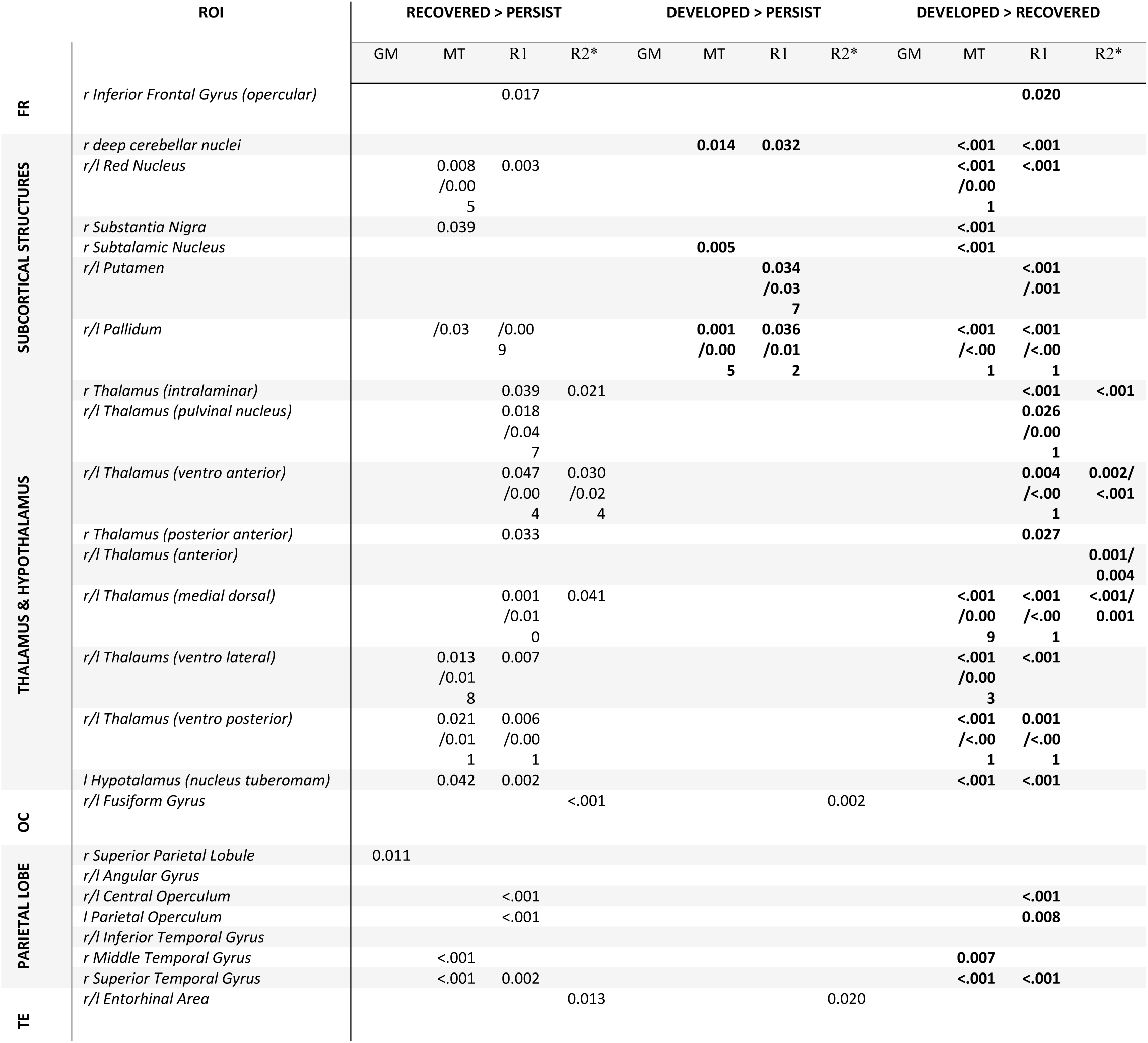
p-values from the Bonferroni corrected linear model where the four trajectories of CP in time were regressed against each ROI individually, for each biophysical model (GM, MT, R1, R2s) with age, gender and sum of compound of interest (or TIV) as covariates of non interest. Each row represents one ROI and each column one of the models for the pair of groups compared. The tables show three group comparisons each, parameter in bold are the positive effects in terms of difference between the two groups indicated on the top of the columns. Only significant effects are shown.

## References

[1] Alexander AL, Hurley SA, Samsonov AA, Adluru N, Hosseinbor AP, Mossahebi P, Tromp DPM, Zakszewski E, Field AS. Characterization of Cerebral White Matter Properties Using Quantitative Magnetic Resonance Imaging Stains. Brain Connectivity 2011;1:423–446.

[2] Balbastre Y, Aghaeifar A, Corbin N, Brudfors M, Ashburner J, Callaghan MF. Correcting inter-scan motion artifacts in quantitative R_1_ mapping at 7T. Magnetic Resonance in Med 2022;88:280– 291.

[3] Bouhassira D, Lantéri-Minet M, Attal N, Laurent B, Touboul C. Prevalence of chronic pain with neuropathic characteristics in the general population. Pain 2008;136:380–387.

[4] Bridges AJ, Gregus SJ, Rodriguez JH, Andrews AR, Villalobos BT, Pastrana FA, Cavell TA. Diagnoses, intervention strategies, and rates of functional improvement in integrated behavioral health care patients. Journal of Consulting and Clinical Psychology 2015;83:590– 601.

[5] Bussy A, Patel R, Parent O, Salaciak A, Bedford SA, Farzin S, Tullo S, Picard C, Villeneuve S, Poirier J, Breitner JC, Devenyi GA, Tardif CL, Chakravarty MM. Exploring morphological and microstructural signatures across the Alzheimer’s spectrum and risk factors. Neurobiology of Aging 2025;149:1–18.

[6] Castella R, Arn L, Dupuis E, Callaghan MF, Draganski B, Lutti A. Controlling motion artefact levels in MR images by suspending data acquisition during periods of head motion. Magnetic Resonance in Med 2018;80:2415–2426.

[7] Cheli VT, Correale J, Paez PM, Pasquini JM. Iron Metabolism in Oligodendrocytes and Astrocytes, Implications for Myelination and Remyelination. ASN Neuro 2020;12. doi:10.1177/1759091420962681.

[8] Chen T, Pei X, Bai X, Zhang X, Zhao Y, Lin C, Yang Q, Sun S, Wang Y, Sui B. Abnormal brain iron deposition in patients with new daily persistent headache: a prospective quantitative susceptibility mapping study. J Headache Pain 2025;26:239.

[9] Conboy V, Edwards C, Ainsworth R, Natusch D, Burcham C, Danisment B, Khot S, Seymour R, Larcombe SJ, Tracey I, Kolasinski J. Chronic musculoskeletal impairment is associated with alterations in brain regions responsible for the production and perception of movement. The Journal of Physiology 2021;599:2255–2272.

[10] Corbin N, Oliveira R, Raynaud Q, Di Domenicantonio G, Draganski B, Kherif F, Callaghan MF, Lutti A. Statistical analyses of motion-corrupted MRI relaxometry data computed from multiple scans. Journal of Neuroscience Methods 2023;398:109950.

[11] Cuyul-Vásquez I, Ponce-Fuentes F, Salazar J, Fuentes J, Araya-Quintanilla F. Can exercise-based interventions reverse gray and white matter abnormalities in patients with chronic musculoskeletal pain? A systematic review. BMR 2023;36:957–968.

[12] Davis KD, Moayedi M. Central Mechanisms of Pain Revealed Through Functional and Structural MRI. J Neuroimmune Pharmacol 2013;8:518–534.

[13] De Zoete RMJ, Berryman CF, Nijs J, Walls A, Jenkinson M. Differential Structural Brain Changes Between Responders and Nonresponders After Physical Exercise Therapy for Chronic Nonspecific Neck Pain. The Clinical Journal of Pain 2023;39:270–277.

[14] Dirupo G, Rossel J, Fournier N, D’Andrea A, Vollenweider P, Decosterd I, Suter MR, Berna C. Correlates of chronic pain onset and recovery in the CoLaus cohort. European Journal of Pain 2024;29. doi:10.1002/ejp.4712.

[15] Domin M, Grimm NK, Klepzig K, Schmidt CO, Kordass B, Lotze M. Gray Matter Brain Alterations in Temporomandibular Disorder Tested in a Population Cohort and Three Clinical Samples. The Journal of Pain 2021;22:739–747.

[16] Draganski B, Ashburner J, Hutton C, Kherif F, Frackowiak RSJ, Helms G, Weiskopf N. Regional specificity of MRI contrast parameter changes in normal ageing revealed by voxel-based quantification (VBQ). NeuroImage 2011;55:1423–1434.

[17] Fairhurst M, Wiech K, Dunckley P, Tracey I. Anticipatory brainstem activity predicts neural processing of pain in humans. Pain 2007;128:101–110.

[18] Firmann M, Mayor V, Vidal PM, Bochud M, Pécoud A, Hayoz D, Paccaud F, Preisig M, Song KS, Yuan X, Danoff TM, Stirnadel HA, Waterworth D, Mooser V, Waeber G, Vollenweider P. The CoLaus study: a population-based study to investigate the epidemiology and genetic determinants of cardiovascular risk factors and metabolic syndrome. BMC Cardiovasc Disord 2008;8. doi:10.1186/1471-2261-8-6.

[19] Gagnon CM, Scholten P, Atchison J, Jabakhanji R, Wakaizumi K, Baliki M. Structural MRI Analysis of Chronic Pain Patients Following Interdisciplinary Treatment Shows Changes in Brain Volume and Opiate-Dependent Reorganization of the Amygdala and Hippocampus. Pain Medicine 2020;21:2765–2776.

[20] Grosu C, Trofimova O, Gholam-Rezaee M, Strippoli M-PF, Kherif F, Lutti A, Preisig M, Draganski B, Eap CB. CYP2C19 expression modulates affective functioning and hippocampal subiculum volume—a large single-center community-dwelling cohort study. Transl Psychiatry 2022;12:316.

[21] Hagen M, Madhavan T, Bell J. Combined analysis of 3 cross-sectional surveys of pain in 14 countries in Europe, the Americas, Australia, and Asia: impact on physical and emotional aspects and quality of life. Scandinavian Journal of Pain 2020;20:575–589.

[22] Hastie CE, Foster HME, Jani BD, O’Donnell CA, Ho FK, Pell JP, Sattar N, Katikireddi SV, Mair FS, Nicholl BI. Chronic pain and COVID-19 hospitalisation and mortality: a UK Biobank cohort study. Pain 2023;164:84–90.

[23] Helms G, Dathe H, Dechent P. Quantitative FLASH MRI at 3T using a rational approximation of the Ernst equation. Magnetic Resonance in Med 2008;59:667–672.

[24] Helms G, Dathe H, Kallenberg K, Dechent P. High-resolution maps of magnetization transfer with inherent correction for RF inhomogeneity and T_1_ relaxation obtained from 3D FLASH MRI. Magnetic Resonance in Med 2008;60:1396–1407.

[25] Helms G, Dechent P. Increased SNR and reduced distortions by averaging multiple gradient echo signals in 3D FLASH imaging of the human brain at 3T. Magnetic Resonance Imaging 2009;29:198–204.

[26] Johnston KJA, Adams MJ, Nicholl BI, Ward J, Strawbridge RJ, Ferguson A, McIntosh AM, Bailey MES, Smith DJ. Genome-wide association study of multisite chronic pain in UK Biobank. PLoS Genet 2019;15:e1008164.

[27] Keuken MC, Bazin P-L, Backhouse K, Beekhuizen S, Himmer L, Kandola A, Lafeber JJ, Prochazkova L, Trutti A, Schäfer A, Turner R, Forstmann BU. Effects of aging on T1, T∗2, and QSM MRI values in the subcortex. Brain Struct Funct 2017;222:2487–2505.

[28] Kuner R, Flor H. Structural plasticity and reorganisation in chronic pain. Nat Rev Neurosci 2017;18:20–30.

[29] Lam J, Mårtensson J, Westergren H, Svensson P, Sundgren PC, Alstergren P. Structural MRI findings in the brain related to pain distribution in chronic overlapping pain conditions: An explorative case–control study in females with fibromyalgia, temporomandibular disorder-related chronic pain and pain-free controls. J of Oral Rehabilitation 2024;51:2415– 2426.

[30] Lambert C, Simon H, Colman J, Barrick TR. Defining thalamic nuclei and topographic connectivity gradients in vivo. NeuroImage 2017;158:466–479.

[31] Liao X, Mao C, Wang Y, Zhang Q, Cao D, Seminowicz DA, Zhang M, Yang X. Brain gray matter alterations in Chinese patients with chronic knee osteoarthritis pain based on voxel-based morphometry. Medicine 2018;97:e0145.

[32] Loggia ML. “Neuroinflammation”: does it have a role in chronic pain? Evidence from human imaging. Pain 2024;165:S58–S67.

[33] Lorio S, Kherif F, Ruef A, Melie-Garcia L, Frackowiak R, Ashburner J, Helms G, Lutti A, Draganski B. Neurobiological origin of spurious brain morphological changes: A quantitative MRI study. Human Brain Mapping 2016;37:1801–1815.

[34] Loued-Khenissi L, Trofimova O, Vollenweider P, Marques-Vidal P, Preisig M, Lutti A, Kliegel M, Sandi C, Kherif F, Stringhini S, Draganski B. Signatures of life course socioeconomic conditions in brain anatomy. Human Brain Mapping 2022;43:2582–2606.

[35] Lutti A, Corbin N, Ashburner J, Ziegler G, Draganski B, Phillips C, Kherif F, Callaghan MF, Di Domenicantonio G. Restoring statistical validity in group analyses of motion-corrupted MRI data. Human Brain Mapping 2022;43:1973–1983.

[36] Lutti A, Hutton C, Finsterbusch J, Helms G, Weiskopf N. Optimization and validation of methods for mapping of the radiofrequency transmit field at 3T. Magnetic Resonance in Med 2010;64:229–238.

[37] Lutti A, Stadler J, Josephs O, Windischberger C, Speck O, Bernarding J, Hutton C, Weiskopf N. Robust and Fast Whole Brain Mapping of the RF Transmit Field B1 at 7T. PLoS ONE 2012;7:e32379.

[38] Macfarlane GJ, Barnish MS, Jones GT. Persons with chronic widespread pain experience excess mortality: longitudinal results from UK Biobank and meta-analysis. Annals of the Rheumatic Diseases 2017;76:1815–1822.

[39] May A. Chronic pain may change the structure of the brain. Pain 2008;137:7–15.

[40] McCarberg B, Peppin J. Pain Pathways and Nervous System Plasticity: Learning and Memory in Pain. Pain Medicine 2019;20:2421–2437.

[41] Medrano-Escalada Y, Plaza-Manzano G, Fernández-de-las-Peñas C, Valera-Calero JA. Structural, Functional and Neurochemical Cortical Brain Changes Associated with Chronic Low Back Pain. Tomography 2022;8:2153–2163.

[42] Mosch B, Hagena V, Herpertz S, Diers M. Brain morphometric changes in fibromyalgia and the impact of psychometric and clinical factors: a volumetric and diffusion-tensor imaging study. Arthritis Res Ther 2023;25:81.

[43] Neumann N, Domin M, Lotze M. Gray matter volume of limbic brain structures during the development of chronic back pain: a longitudinal cohort study. Pain 2025;166:438–447.

[44] Noorani A, Hung PS-P, Zhang JY, Sohng K, Laperriere N, Moayedi M, Hodaie M. Pain Relief Reverses Hippocampal Abnormalities in Trigeminal Neuralgia. The Journal of Pain 2022;23:141–155.

[45] Ortega N, Mueller NJ, Dehghan A, De Crom TOE, Von Gunten A, Preisig M, Marques-Vidal P, Vinceti M, Voortman T, Rodondi N, Chocano-Bedoya PO. Dairy intake and cognitive function in older adults in three cohorts: a mendelian randomization study. Nutr J 2025;24:20.

[46] Ortiz E, Pasquini JM, Thompson K, Felt B, Butkus G, Beard J, Connor JR. Effect of manipulation of iron storage, transport, or availability on myelin composition and brain iron content in three different animal models. J of Neuroscience Research 2004;77:681–689.

[47] Pan PL, Zhong JG, Shang HF, Zhu YL, Xiao PR, Dai ZY, Shi HC. Quantitative meta-analysis of grey matter anomalies in neuropathic pain. European Journal of Pain 2015;19:1224–1231.

[48] Pashkov A, Filimonova E, Zaitsev B, Martirosyan A, Moysak G, Rzaev J. Thalamic changes in patients with chronic facial pain. Neuroradiology 2025;67:895–908.

[49] Piñero DJ, Connor JR. Iron in the Brain: An Important Contributor in Normal and Diseased States. Neuroscientist 2000;6:435–453.

[50] Pomares FB, Funck T, Feier NA, Roy S, Daigle-Martel A, Ceko M, Narayanan S, Araujo D, Thiel A, Stikov N, Fitzcharles M-A, Schweinhardt P. Histological Underpinnings of Grey Matter Changes in Fibromyalgia Investigated Using Multimodal Brain Imaging. J Neurosci 2017;37:1090–1101.

[51] Preisig M, Waeber G, Vollenweider P, Bovet P, Rothen S, Vandeleur C, Guex P, Middleton L, Waterworth D, Mooser V, Tozzi F, Muglia P. The PsyCoLaus study: methodology and characteristics of the sample of a population-based survey on psychiatric disorders and their association with genetic and cardiovascular risk factors. BMC Psychiatry 2009;9. doi:10.1186/1471-244x-9-9.

[52] Reckziegel D, Abdullah T, Wu B, Wu B, Huang, L, Schnitzer T, Apkarian V. Hippocampus shape deformation: a potential diagnostic biomarker for chronic back pain in women. PAIN 2021;162:1457–1467.

[53] Rönnegård A-S, Nowak C, Äng B, Ärnlöv J. The association between short-term, chronic localized and chronic widespread pain and risk for cardiovascular disease in the UK Biobank. European Journal of Preventive Cardiology 2022;29:1994–2002.

[54] Salami A, Papenberg G, Sitnikov R, Laukka EJ, Persson J, Kalpouzos G. Elevated neuroinflammation contributes to the deleterious impact of iron overload on brain function in aging. NeuroImage 2021;230:117792.

[55] Tabelow K, Balteau E, Ashburner J, Callaghan MF, Draganski B, Helms G, Kherif F, Leutritz T, Lutti A, Phillips C, Reimer E, Ruthotto L, Seif M, Weiskopf N, Ziegler G, Mohammadi S. hMRI – A toolbox for quantitative MRI in neuroscience and clinical research. NeuroImage 2019;194:191–210.

[56] Todorich B, Pasquini JM, Garcia CI, Paez PM, Connor JR. Oligodendrocytes and myelination: The role of iron. Glia 2009;57:467–478.

[57] Trofimova O, Loued-Khenissi L, DiDomenicantonio G, Lutti A, Kliegel M, Stringhini S, Marques-Vidal P, Vollenweider P, Waeber G, Preisig M, Kherif F, Draganski B. Brain tissue properties link cardio-vascular risk factors, mood and cognitive performance in the CoLaus|PsyCoLaus epidemiological cohort. Neurobiology of Aging 2021;102:50–63.

[58] Urrutia PJ, Bórquez DA, Núñez MT. Inflaming the Brain with Iron. Antioxidants 2021;10:61.

[59] Van Grinsven M, Witkam R, Kurt E, Özkan S, Van Der Kolk A, Vissers K, Henssen D. Thalamic Microstructural Alterations as Revealed by the T1/T2 Ratio in Chronic Pain Patients. JCM 2025;14:2888.

[60] Vos De Wael R, Larivière S, Caldairou B, Hong S-J, Margulies DS, Jefferies E, Bernasconi A, Smallwood J, Bernasconi N, Bernhardt BC. Anatomical and microstructural determinants of hippocampal subfield functional connectome embedding. Proc Natl Acad Sci USA 2018;115:10154–10159.

[61] Wang Z, Yuan M, Xiao J, Chen L, Guo X, Dou Y, Jiang F, Min W, Zhou, B. Gray Matter Abnormalities in Patients with Chronic Primary Pain: A Coordinate-Based Meta-Analysis. Pain Physician 2022;25:1–13.

[62] Ward RJ, Zucca FA, Duyn JH, Crichton RR, Zecca L. The role of iron in brain ageing and neurodegenerative disorders. The Lancet Neurology 2014;13:1045–1060.

[63] World Health Organization. ICD-11: International Classification of Diseases (11th Revision). 2022. Available: https://icd.who.int/.

[64] Yao D, Chen Y, Chen G. The role of pain modulation pathway and related brain regions in pain. Reviews in the Neurosciences 2023;34:899–914.

[65] Zhang Y, Liu F-Y, Liao F-F, Wan Y, Yi M. Exacerbation of tonic but not phasic pain by entorhinal cortex lesions. Neuroscience Letters 2014;581:137–142.

[66] Zorina-Lichtenwalter K, Bango C, Cecko M, Reader L, Lindquist M, Friedman N, Wager T. Brain Correlates of Chronic Pain Onset, Progression, and Resolution. The Journal of Pain 2024;25:46.

